# Mothers’ hygiene experiences in confinement centres: a cohort study

**DOI:** 10.1101/2021.06.28.21259614

**Authors:** Siew Cheng Foong, Wai Cheng Foong, May Loong Tan, Jacqueline Judith Ho

## Abstract

Ethnic Malaysian Chinese used to observe the 1-month postpartum confinement period at home and many families would engage a traditional postpartum carer (TPC) to help care for the mother and newborn. A recent trend has been the development of confinement centres (CCs) which are private non-healthcare establishments run by staff not trained in health care. Concerns about hygiene in CCs arose after infections were reported. We describe the practice of hand hygiene observed in CCs, the availability of resources for hygiene and to determine the prevalence of health related problems in CCs.

**Methods:** This is a cohort study of ethnic Chinese mothers intending to breastfeed their healthy infants. They were recruited post-delivery along with a comparison group who planned to spend their confinement period at home, then all were telephone interviewed after their 1-month confinement period about their experience. To avoid any alteration in behaviour, mothers were not told at recruitment that they had to observe hygiene practices. Multiple logistic regression was used to assess the effect of place of confinement on rates of infant health problems.

**Results:** Of 187 mothers, 88 (47%) went to 27 different CCs while 99 (53%) stayed at home. Response rates for the 1-month interviews were 88% (CC) versus 97% (home).

Mothers in CC group stayed in one to four-bedded rooms and 92% of them had their baby sleeping separately in a common nursery described to have up to 17 babies at a time; 74% of them spent less than six hours a day with their babies; 43% noticed that CC staff had inadequate hand hygiene practices; 66% reported no hand-basins in their rooms; 30% reported no soap at hand-basins; 28% reported inexperienced or inadequate staff and 4% reported baby item sharing.

Of mothers staying at home, 35% employed a TPC to care for her baby; 32% did not room-in with their babies, but only 11% spent less than 6 hours a day with their babies. 18% of mothers who employed TPCs reported that their TPC had unsatisfactory hand hygiene practices.

Health problems that were probably related to infection (HPRI) like fever and cough were similar between the groups: 14% (CC) versus 14% (home) (p=0.86). Multiple logistic regression did not show that CCs were a factor for HPRI: aOR 1.28 (95% CI 0.36 to 4.49). Three mothers reported events that could indicate transmission of infection in CCs.

**Conclusion:** We found unsatisfactory hygiene practices in CCs as reported by mothers who spent their confinement period there. Although we were not able to establish any direct evidence of infection transmission but based on reports given by the mothers in this study, it is likely to be happening. Therefore, future studies, including intervention studies, are urgently needed to establish an appropriate hygiene standard in CCs as well as the best method to implement this standard. Empowering CC staff with hygiene knowledge so that they can be involved and contribute to the development of the development of these standards would be important.

## Background

The 30-day postpartum (confinement) period is an important time for women of Chinese ethnicity in Malaysia and elsewhere [1, 2]. Despite modernisation, most families still adhere strictly to traditional practices based on maintaining the Yin and Yang with the belief that failure to do so could potentially be detrimental to the mother’s health [2-5]. Many families engaged a traditional postpartum carer (TPC), known locally as a ‘confinement lady’ or ‘yue sao’ to stay in the new mother’s home during the confinement period and assist the mother. The TPC is traditionally someone who is considered an expert in the necessary postpartum diet and practices. Their skills were probably obtained through experience rather than formal training [1, 6].

Over the last decade, confinement centres (CCs) where post-partum Chinese mothers could stay and observe traditional post-partum practices during their confinement period, have emerged as an alternative option. CCs are private establishments, usually converted from residential or commercial properties, with rooms for mothers’ accommodation. CC staff are generally women who are familiar with the Chinese cultural confinement requirements and diet, similar to a TPC. Although some CCs do employ qualified nurses and midwives [6], others may employ untrained staff to help [7].

Concerns about hygiene in these centres arose when anecdotal reports suggested that babies in CCs were frequently hospitalised with serious infections. In 2007, Rai et al published a report about an Echovirus infection outbreak in a CC and poor hygiene practices in that CC were highlighted [8]. Therefore, to learn more about how hygiene is practiced in CCs, we asked mothers who had chosen to stay in a confinement centre about hygiene practices they had observed during the stay. Mothers who had employed a TPC to help them at home were also asked about the TPC’s hygiene practice. This is part of a larger study where we looked at mothers’ breastfeeding experiences in CCs and compared these with a cohort of women who had their traditional confinement period at home [7].

The aim of this paper is to describe the practice of hand hygiene observed in CCs, the availability of resources for hygiene and to determine the prevalence of health related problems in CCs.

## Methods

Details of the study methods are published in Foong 2021 [7] and we describe these in brief here. Malaysian mothers of Chinese ethnicity who intended to spend their confinement period in a CC, who had delivered term healthy infants and had the intention to breastfeed, were recruited prior to discharge. We also recruited a comparison group of mothers who went home for their confinement period, some of whom engaged a TPC. For this paper we used this comparison group to gauge hand hygiene practices in the CC with the TPC and to compare health-related problems in CCs with those in the community. Recruitment and collection of baseline data was done by the baby’s attending doctor, who apart from this was not otherwise involved in the study. Written consent was obtained from the mothers prior to recruitment. To avoid any alteration in behaviour, mothers were not told at recruitment that they would be asked about hygiene practices.

After discharge, there was no contact between the research team and the mother until immediately after her 30-day confinement period. At this point we conducted a telephone interview with all mothers. We firstly asked mothers questions related to their baby’s general health. We then categorised the reported health related problems to those that we judged were possibly related to infection and those that were probably not related to infections. Health problems possibly related to infection (HPRI) included fever, diarrhoea and baby being inactive. Those that we judged to be health problems that were unlikely related to infection (HPUI) included neonatal jaundice and regurgitation of feeds. If any health problem was reported, we asked if they had sought advice from a healthcare professional.

Where applicable, we asked whether or not they had observed their CC staff or TPC (in the case of those at home), washing their hands before handling their babies; and their response if hand-washing practices were not observed. We did not ask mothers staying at home who did not employ TPC whether or not their family members washed hands because the information was deemed to be possibly unreliable. Specifically for mothers who went to CCs, we asked the number of rooms in their CC, the number of mothers staying together in a room, if they had a hand basin in their room, if soap was available in all hand basins, what was provided for mothers to dry their hands, whether alcohol hand sanitizers were available, the number of babies in each nursery, availability of quarantine rooms for sick mothers or babies and their perception of cleanliness in their CC (either very dirty, somewhat dirty, clean or very clean). At the end of the interview, we asked mothers to share with us anything else they would like to about their experience in CCs.

All questions used had been tested in a separate group of mothers not involved in the study. The telephone interview was conducted by three trained research staff and the responses were directly entered into a specially designed interview form. A sample size of 188 was calculated based on the primary objectives of the primary study. Details of this are published in Foong 2021 [7]. This study received ethical approval from the Joint Penang Independent Ethics Committee (JPEC No 08-17-0103).

### Data analyses

We tabulated the baseline demographics of the mothers according to place of confinement. Continuous data was presented as means with standard deviations (SD) and categorical data presented as frequency with percentage (%). Chi-square analysis was used to compare the baseline characteristics between mothers staying in confinement centres (CCs) and those staying at home. Comments from free field options were tabulated and categorized into groups. Some of these free field responses were quoted as illustrations. We used simple logistic regression; and multiple logistic regression after adjusting for clinically important confounders to determine the likelihood of HPRI and HPUI as a function of CCs. The results were presented as crude and adjusted odd ratios (cOR and aOR) with 95% confidence intervals (CI). Statistical analysis was done using Stata 13 [9]. We considered a p-value of less than 0.05 as significant.

## Results

A total of 187 mothers consented to participate, of which 88 (47%) stayed in a CC and 99 (53%) went home. At one month post-partum we were able to interview 77 (88%) mothers from the CC group and 96 (97%) from the home group. Based on the reported names of the CCs given by the mother, the 77 mothers in the CC group had gone to one of probably 27 CCs during their confinement period. Unfortunately we were not able to verify reported names of CC because at the time of the study there was no record of all CCs in Penang available and we are therefore uncertain about the exact number of CCs in the study. Of the 96 mothers from the home group, 34 hired a TPC while the remainder received care from family members. (Figure 1).

**Figure 1:**
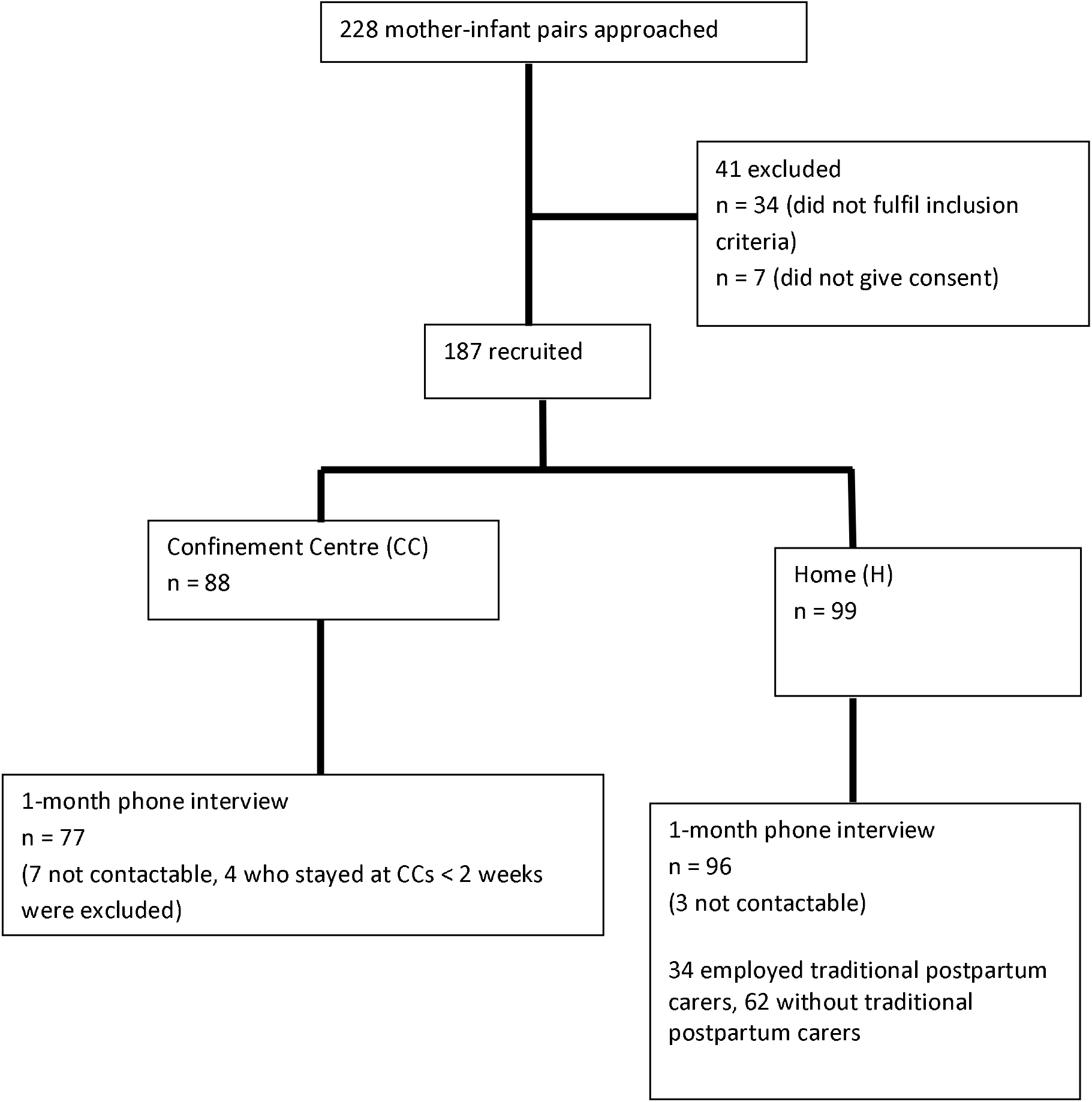
Study flow diagram.

### Baseline characteristics

The maternal and infant baseline characteristics are presented in Table 1. The overall mean maternal age was 32 (SD 4) years. Most mothers had tertiary education, and all had at least secondary school education, which reflects what is expected in Penang. The overall mean infant gestational age was 39 (SD 1) weeks and mean birth weight 3149 (SD 322) g. We found that significantly more primiparas went to CCs (53% CC vs 34% H, p = 0.01) but there were no differences in the age, education background, mode of delivery; infant gestation and birth weight between the two groups. (Table 1).

**Table 1:** Baseline characteristics of the mothers and infants (n = 187)

| Characteristics | Place of confinement, n (%) |  |
| --- | --- | --- |
|  | Confinement centre<br>(n = 88) | Home<br>(n = 99) |
| Age of mothers (years), mean (SD) | 32 (4) | 32 (3) |
| Received tertiary Education | 70 (80) | 80 (81) |
| Primigravida* | 47 (53) | 34 (34) |
| Male infant | 45 (51) | 56 (57) |
| Gestational age at birth (weeks), mean (SD) | 39 (1) | 39 (1) |
| Infant's birth weight (g), mean (SD) | 3141 (304) | 3156 (339) |

\* $p$ value < 0.05

### Description of confinement centres

The description of the CCs came from mother’s reports during the interview. More than one mother may have stayed in the same CCs. The CCs had between four to 10 rooms for mother’s accommodation.

The number of mothers staying together in a room ranged from one to four. 45 mothers occupied a single bedded room, 20 mothers occupied a two bedded room, 11 occupied three bedded room and one stayed in a four bedded room. Most of the mothers did not room-in with their babies beside them (n = 71, 92%). Instead, their babies slept in the common nursery; and majority of mothers (n = 57, 74%) spent less than six hours a day with their babies. Regardless of CC size, all had a single common nursery for babies. The number of babies in the nursery at a time was reported to range from one to 17.

Of the mothers staying at home, 31 (32%) did not room-in with their babies, but only 11 (11%) spent less than six hours a day with their babies.

### Hygiene and infection control measures at CC

When asked to rate the overall cleanliness of the CCs, all mothers reported that their centre was either ‘clean’ (n = 41, 53%) or ‘very clean’ (n = 36, 47%). However, only 17 (22%) mothers noticed that their CC staff washed hands in between handling babies and 33 (43%) mothers noticed that CC staff did not. When asked what they did if the CC staff failed to wash hands before handling a baby, two mothers reported that they went on to ask the staff to do so; two mothers said that not washing hands in between handling babies was not of any concern, and one just said that she felt sorry for the staff who was short-handed at that time. The remaining 27 (35%) mothers had not paid attention to whether or not their CC staff washed hands. (Table 2).

**Table 2:** Mothers’ perception that hand hygiene was practiced before handling babies.

|  | Hand hygiene practised | Hand hygiene not practised | Don't know if hand hygiene practised |
| --- | --- | --- | --- |
| CC staff (n, %) | 17 (22) | 33 (43) | 27 (35) |
| TPC (n, %) | 34 (35) | 6 (18) | 11 (32) |

Only 55 (63%) of mothers reported that their CCs supplied hand soap. Among the 32 (36%) mothers who reported that their CCs provided alcohol-based hand sanitizers, three reported that alcohol-based hand sanitizers were restricted to staff use only. Twenty-six (34%) mothers reported the availability of a sink for hand washing in their room. Only 23 mothers reported availability of hand towels for drying hands and these items were reported to be either a single cloth-towel that was shared by everyone in the centre or toilet rolls. (Table 3).

**Table 3:** Mothers’ perception of the availability of hand hygiene resources at CCs (n = 77)

| Hand hygiene resource | Available (n) |
| --- | --- |
| Sink in own room | 26 |
| Readily available hand soap at each sink | 55 |
| Hand towels to dry hands | 23 |
| Readily available alcohol hand sanitizers | 32 |

The availability of a quarantine room for sick mothers was reported by 17% of mothers while the availability of quarantine rooms for sick babies were reported by 24% of mothers.

One mother reported that her CC required all visitors to don gowns prior to entering the nursery. During the course of the interview, mothers also revealed one or more of the comments related to poor hygiene listed in Table 4.

**Table 4:** Comments related to hygiene in CCs.

|  |
| --- |
| Staff shortage and inexperienced staff who were unaware of hygiene practices |
| Only one toilet to be shared by all mothers hence quite dirty |
| A common towel used to burp all babies in the nursery |
| A common hand towel used by all mothers to wipe their hands |
| The same pail that was used for holding bath water was used for washing the floor |
| The same basin used to wash babies' bottoms was also used to wash milk bottles |
| Milk bottles that fell to the floor (staff fell asleep) were simply picked up and used to continue feeding the baby without being washed |
| Breast-pump parts were just soaked in hot water and not properly sterilized |
| Use of a common milk bottle that was sticky and dirty looking |
| Infrequent changing of diapers, cot sheets and bed sheets |
| Nursery cramped and not spaced |
| 3 babies sharing a single cot |
| Flies and mosquitos in their room |
| Poor quality paper hand towels - toilet rolls that easily disintegrate |
| Alcohol hand sanitizers were only for staff usage |

With regards to hygiene practices by TPCs at home, 6 (18%) of mother reported that their TPC did not washed hands before handling their baby and after changing diapers while 11 (32%) mothers did not know whether their TPC practiced hand hygiene. When we asked what they did when they saw poor hand hygiene, two reported that they asked their TPC to do so, while four did not do anything.

### Babies’ general health at CCs and at home

Baby’s general health at CCs and home were generally similar. HPRI were reported by 11 mothers (14%) from CCs compared to 13 mothers (14%) from home; p value 0.86. Of these, 10 mothers from CC compared to 13 mothers from home consulted a healthcare professional. Reported HPRI included one or more of these: ‘fever’, ‘viral infection’, ‘cough’, ‘stuffy nose’ ‘runny nose’ and ‘oral thrush’. None of the infants from the CC group had any form of serious illness. One infant from the Home group was hospitalised for viral infection which the mother thought was caught from the baby’s older brother.

The main HPUI was ‘jaundice’. Others included one or more of the following: ‘colic’, ‘constipation’, ‘regurgitation’ and ‘umbilical hernia’. HPUI were reported by 70 mothers (92%) from CCs compared to 88 mothers (92%) from home; p value 0.92. Of these, 43 mothers from CC and 63 mothers from home consulted a healthcare professional.

Simple logistic regression showed no association between HPRI and place of confinement, OR 1.08 (95% CI 0.45 to 2.57), p = 0.86. There was also no association between HPUI and place of confinement, OR 1.06 (95% CI 0.35 to 3.20). Multiple logistic regression adjusted for known clinically important confounders (maternal education level, spent less than six hours a day with baby, not sleeping with baby at night and no exclusive breastfeeding) also did not show that the CC or home was a factor affecting HPRI (aOR 1.28 (95% CI (0.36 to 4.49), p = 0.71); or HPUI (aOR 2.01(95% CI 0.52) to 7.82, p=0.32). (Table 5).

**Table 5:** Crude and adjusted ORs for HPRI and HPUI defined by place of confinement.

|  |  | Number of mothers who reported (n) | Odds Ratio, OR (95% CI) | Adjusted Odds Ratio, aOR (95% CI) <sup>a</sup> |
| --- | --- | --- | --- | --- |
| HPRI | Confinement Centres (n = 77) | 11 | 1.08 (0.45, 2.57) | 1.28 (0.36, 4.49) |
|  | Home (n = 96) | 13 |  |  |
| HPUI | Confinement Centres (n = 77) | 70 | 1.06 (0.35, 3.20) | 2.01 (0.52, 7.82) |
|  | Home (n = 96) | 88 |  |  |

<sup>a</sup> Adjusted for maternal education level, spent less than six hours a day with baby, not sleeping with baby at night and no exclusive breastfeeding
HPRI: Health problems probably related to infections
HPUI: Health problems probably unrelated to infections

When we asked mothers if they had anything else to share with us, we found three events that could indicate a possibility of infection transmission in CCs. One mother reported that there were visits from the health authorities to her CC because a number of babies in her CC had fever and were hospitalized. One mother reported that all babies in her CC had either blocked or runny noses within a one-week period. Another thought her baby’s oral thrush and rashes on her baby were due to sharing of a baby wash cloth at the CC. The mothers who reported these events probably came from three different CCs.

## Discussion

The main finding from our study was inadequate hand hygiene among caregivers of babies not only in CCs but also at home. We also found inadequate infection control facilities in CCs. Despite this, mothers reported that they were satisfied with the cleanliness of their CCs. Due to small numbers, we were unable to show whether there was a difference for the type and prevalence of reported health problems between the two groups. However, practices at CCs that may potentially have caused infection were noted. These practices most likely result from lack of resources, inconsistent hand hygiene practices and over-crowding, and perhaps lack of awareness.

We are unable to find other studies looking at confinement centres, but our study draws parallels with studies conducted with nursing homes and child care centres. One similarity with these is that they are populations who are relatively susceptible to infection. These studies found that over-crowding and lack of hand hygiene led to infection transmission [10, 11]. A number of studies describe how infection could be prevented through improving hand hygiene practices, the availability of resources and improved role modelling [12-16]. In addition, these studies also found that education and training could effectively increase hygiene practices in nursing homes [11, 16, 17]. Drawing from the findings of these studies, it is very likely that all of these could apply to CCs. Therefore, we could expect that if education and training were put in place, hygiene practices in CCs could improve. However, good hand hygiene practices are known to be one of those practices that are difficult to sustain and therefore regular audit and feedback to improve sustainability might also be needed[18].

Current guidelines for hygiene practice in healthcare settings differ little in their recommendations but little is known about the appropriate standard of care in community settings such as CCs. Infection control as it is practiced in healthcare settings may be difficult to implement in CCs and is costly. There is currently little research to guide practice. It is noted that CCs are not healthcare institutions, and their staff are not healthcare staff. In addition, the traditional confinement care offered by CCs is not a medical treatment but at the same time CCs need to be cognisant of the increased infection risk of neonates and have adequate infection prevention strategies in place.

Studies have shown that nursing homes struggle to strike a balance in attempting to preserve a homelike environment and hospital-level measures to control of infection [11, 19, 20]. This is likely to apply to the CC environment as well. CCs would need to consider what measures if implemented would be accepted by both staff and mothers and could be complied with. However, at the same time there is no doubt that infection control measures are needed and hand hygiene is obviously the place to start. Research in this area is much needed as well as research on effective training and methods of consolidating hand hygiene practice in CCs.

Many of the home-based TPCs in our control group were also reported to not practice good hand hygiene. There is currently no literature about their hygiene knowledge and practices. However, to improve safe practices, home-based TPCs should also be drawn into training interventions.

We found many mothers who stayed at CCs were discouraged from rooming-in with their babies, and their babies spent most of the time in the nursery [7]. Since there is a body of evidence showing that both mother-infant rooming-in and breastfeeding prevent infection [21-24], ways to improve these practices could also be looked at. Although exclusive breastfeeding rates in this study cohort were as good if not better that other local populations, direct breastfeeding rates were poor [7]. Feeding expressed breastmilk carries an increased risk of infection since it involves use of breast pumps and bottles which need a high level of disinfection [25]. One way to improve direct breastfeeding would be to empower CC staff to provide support for direct breastfeeding and to provide rooming-in facilities for mother and baby. This might mean that maternal accommodation needs further study to establish appropriate recommendations, for example, spacing between mothers. There is also a possibility of considering kangaroo care as a means of infection prevention, but studies are needed both in terms of feasibility and safety.

A limitation of our study would be that we did not have accurate data on which CC the mothers went to. We feel that our sample of mothers reasonably represents the mothers using CCs in Penang, however it is probable not all CCs in Penang were represented in the data. Since our sampling was of women and not CCs and these 77 women went to around 27 CCs, our data represents the number of women and their babies who were exposed to poor hygiene practices and not the number of CCs having poor hygiene practices. Another limitation of the study was the sample size which was not calculated to show a difference in HPRI rates. It is also important to note that our findings are those perceived by mothers. The data was collected after they had left the CC and we did not verify their reports.

Therefore, they could be subject to recall bias as well as observer bias. To overcome this bias, further studies in this area should involve CC operators and managers. Therefore, there is a need to establish rapport with them early.

## Conclusion

We found unsatisfactory hygiene practices in CCs as reported by mothers who spent their confinement period there. Although we were not able to establish any direct evidence of infection transmission but based on reports given by the mothers in this study, it is likely to be happening. Therefore, future studies, including intervention studies, are urgently needed to establish an appropriate hygiene standard in community postpartum care facilities such as these as well as the best method to implement this standard. Empowering CC staff with hygiene knowledge so that they can be involved and contribute to the development of the development of these standards would be important.

## Data Availability

Data are fully available and without restriction.(Contact Vice Dean of Research, RCSI & UCD Malaysia Campus)

## List of abbreviations

CC: Confinement centre
TPC: Traditional postpartum carer
HPRI: Health problems that are probably related to infections
HPUI: Health problems that are unlikely related to infections

## Declarations

### Ethics approval and consent to participate

This study was registered with the National Medical Research Registry (NMRR-17-1174-36384 S1) and ethical approval by Joint Penang Independent Ethics Committee (JPEC 02-18-0026). All mothers gave written informed consent.

### Consent for publication

Not applicable

### Availability of data and material

The datasets used and/or analysed during the current study are available from the corresponding author on reasonable request.

### Authors’ contributions

SCF, WCF, MLT, and JJH were major contributors in the design of the study and in writing the manuscript. WCF and SCF analysed and interpreted the data. All authors read and approved the final manuscript.

## Acknowledgements

The authors acknowledge Ru Jian Jonathan Teoh (initial draft of the protocol); Adele Tan, Hon Kit Cheang, Yee Chern Hwang, Giap Liang Dan, Jessica Tan, Kwai Meng Pong, Siti Khadijah Hamdan, Balkees Abdul Majeed (mother recruitment); Caryn Lim, Wei Wen Lee, Zcho Huey Lee, Claire Lee and Bee Hong Ang (data management); as well as all mothers and hospitals who participated in this study.

## Authors’ information

SCF, MLT, WCF and JJH are all paediatricians and advocates for breastfeeding.

## References

1. Fok D, Aris IM, Ho J, Lim SB, Chua MC, Pang WW, et al. A Comparison of Practices During the Confinement Period among Chinese, Malay, and Indian Mothers in Singapore. Birth. 2016;43(3):247–54.

2. Raven JH, Chen Q, Tolhurst RJ, Garner P. Traditional beliefs and practices in the postpartum period in Fujian Province, China: a qualitative study. BMC pregnancy and childbirth. 2007;7:8.

3. Dennis CL, Fung K, Grigoriadis S, Robinson GE, Romans S, Ross L. Traditional postpartum practices and rituals: a qualitative systematic review. Womens Health (Lond). 2007;3(4):487–502.

4. Ding G, Tian Y, Yu J, Vinturache A. Cultural postpartum practices of ‘doing the month’ in China. Perspect Public Health. 2018;138(3):147–9.

5. Poh B, Koon W, Yuen P, Norimah A. Postpartum dietary intakes and food taboos among Chinese women attending maternal and child health clinics and maternity hospital, Kuala Lumpur. Mal J Nutr. 2005;11:1–21.

6. Su-Lyn B. New mothers paying big bucks to be ‘confined’. Malay Mail. 2013 7 July 2013.

7. Foong SC, Tan ML, Foong WC, Ho JJ, Rahim FF. Comparing breastfeeding experiences between mothers spending the traditional Chinese confinement period in a confinement centre and those staying at home: a cohort study. International breastfeeding journal. 2021;16(1):4.

8. Bina Rai S, Wan Mansor H, Vasantha T, Norizah I, Chua KB. An outbreak of echovirus 11 amongst neonates in a confinement home in Penang, Malaysia. The Medical journal of Malaysia. 2007;62(3):223–6.

9. StataCorp. Stata Statistical Software: Release 13. College Station, TX: StataCorp LP. 2013.

10. Brown KA, Jones A, Daneman N, Chan AK, Schwartz KL, Garber GE, et al. Association Between Nursing Home Crowding and COVID-19 Infection and Mortality in Ontario, Canada. JAMA internal medicine. 2020.

11. Hammerschmidt J, Manser T. Nurses’ knowledge, behaviour and compliance concerning hand hygiene in nursing homes: a cross-sectional mixed-methods study. BMC health services research. 2019;19(1):547.

12. Alp E, Ozturk A, Guven M, Celik I, Doganay M, Voss A. Importance of structured training programs and good role models in hand hygiene in developing countries. Journal of infection and public health. 2011;4(2):80–90.

13. Baldwin A, Mills J, Birks M, Budden L. Role modeling in undergraduate nursing education: an integrative literature review. Nurse education today. 2014;34(6):e18–26.

14. Cure L, Van Enk R. Effect of hand sanitizer location on hand hygiene compliance. American journal of infection control. 2015;43(9):917–21.

15. Perry RN. Role modeling excellence in clinical nursing practice. Nurse education in practice. 2009;9(1):36–44.

16. Yeung WK, Tam WS, Wong TW. Clustered randomized controlled trial of a hand hygiene intervention involving pocket-sized containers of alcohol-based hand rub for the control of infections in long-term care facilities. Infection control and hospital epidemiology. 2011;32(1):67–76.

17. Sax H, Uçkay I, Richet H, Allegranzi B, Pittet D. Determinants of good adherence to hand hygiene among healthcare workers who have extensive exposure to hand hygiene campaigns. Infection control and hospital epidemiology. 2007;28(11):1267–74.

18. Ivers N, Jamtvedt G, Flottorp S, Young JM, Odgaard-Jensen J, French SD, et al. Audit and feedback: effects on professional practice and healthcare outcomes. Cochrane Database of Systematic Reviews. 2012(6).

19. Smith PW, Bennett G, Bradley S, Drinka P, Lautenbach E, Marx J, et al. SHEA/APIC Guideline: Infection prevention and control in the long-term care facility. American journal of infection control. 2008;36(7):504–35.

20. Gould DJ, Moralejo D, Drey N, Chudleigh JH, Taljaard M. Interventions to improve hand hygiene compliance in patient care. Cochrane Database Syst Rev. 2017;9(9):CD005186–CD.

21. Mustajab I, Munir M. A rooming-in program for mothers and newborns at Gunung Wenang General Hospital Manado. Paediatrica Indonesiana. 1986;26(9-10):177–84.

22. Kramer MS, Kakuma R. Optimal duration of exclusive breastfeeding. Cochrane Database of Systematic Reviews. 2012(8).

23. Sankar MJ, Sinha B, Chowdhury R, Bhandari N, Taneja S, Martines J, et al. Optimal breastfeeding practices and infant and child mortality: a systematic review and meta-analysis. Acta paediatrica (Oslo, Norway : 1992). 2015;104(467):3–13.

24. Bowatte G, Tham R, Allen KJ, Tan DJ, Lau M, Dai X, et al. Breastfeeding and childhood acute otitis media: a systematic review and meta-analysis. Acta paediatrica (Oslo, Norway : 1992). 2015;104(467):85–95.

25. Brown SL, Bright RA, Dwyer DE, Foxman B. Breast pump adverse events: reports to the food and drug administration. Journal of human lactation : official journal of International Lactation Consultant Association. 2005;21(2):169–74.

